# Donor race has no role in predicting allograft and patient survival among kidney transplant recipients

**DOI:** 10.1101/2021.04.01.21254772

**Authors:** Kelly Chong, Igor Litvinovich, Shan Shan Chen, Yiliang Zhu, Christos Argyropoulos, Yue-Harn Ng

**Author notes:** **Corresponding author:** Yue-Harn Ng, MD, Box 356174, 1959 NE Pacific Street, Seattle WA 98195.

## Abstract

A heated debate in creatinine-based estimated glomerular filtration rate (eGFR) calculation is the inclusion of race alongside biological factors, such as age and gender. Similarly, the race variable was included in the calculation of the Kidney Donor Risk Index (KDRI) as deceased donor kidneys from black donors have historically been shown to be associated with lower allograft or patient survival. Given the current climate of uncertainty with the use of race in nephrology, we sought to answer the question of whether removing the donor race variable from the KDRI would alter its validity to assess allograft and patient survival. Our modeling and analysis showed that removing donor race from the original KDRI did not alter the overall model predictability of allograft failure or patient mortality. Clinical risk factors included in the KDRI have largely accounted for differential risk between black and other donors. Adding donor race into the KDRI only shifts how risk is attributed to these clinical risk factors, without yielding better prediction of outcomes than the model without race.

---

A heated debate in creatinine-based estimated glomerular filtration rate (eGFR) calculation is the inclusion of race alongside biological factors, such as age and gender.^1-3^ The controversy lies with the race variable (black vs. other) being used as a proxy for muscle mass.^3^ To date, there is insufficient evidence to support that blacks have higher muscle mass than whites.^2-4^ Similarly, the race variable was included in the calculation of the Kidney Donor Risk Index (KDRI) as deceased donor kidneys from black donors have historically been shown to be associated with lower allograft or patient survival.^5^ However, recent evidence suggest that it is not race, but the presence of the APOL1 gene G1 and G2 coding variants that actually confers a worse allograft outcome.^6^ Hence, black race itself does not confer a biological risk for allograft failure and should not be labelled as a risk factor for allograft failure. Given the current climate of uncertainty with the use of race in nephrology, we sought to answer the question of whether removing the donor race variable from the KDRI^7^ would alter its validity to assess allograft and patient survival.

Using the Scientific Registry of Transplant Recipients data from 2000-2017, we included adult patients with kidney transplants only and excluded patients with previous solid organ transplant. We evaluated two proportional hazard survival models for combined outcome of allograft failure or patient death. The first was a replication of the original KDRI model^7^ and the KDRI score was a fixed component of the hazard ratio. In the second model, we removed the donor race variable, refit the remaining components of the hazard ratio, and derived a modified KDRI score without race. We then compared these two models using the area under the curve (AUC) of the Receiver Operating Characteristic curve. We estimated AUC by first randomly selecting 40,000 records as the training sample to fit each of the two models and then used the fitted model to predict outcomes for the remaining patients (n=79,441) at each month over the 17-year study period. We replicated this process 10 times and compared the average AUC of the replications. AUC for survival model is dependent on time-point at which we predict outcome. We used R package survAUC.^8,9^ This study was approved by the institution review board at the University of New Mexico.

We compared donor characteristics between black and other donors (Table 1). Black donors were younger on average, but were not different with respect to height and weight or gender distribution. Black donors were more likely to have diabetes, hypertension or cerebrovascular accident as the cause of death, but less likely to be cardiac death donors or positive for hepatitis C. After removing donor race from KDRI, five donor-related regression coefficients changed less than 1%, four between 1% and 5%, and those of creatinine, HCV, and diabetes changed between 5-11%, corresponding to up to 10% hazard ratio change (see supplement). Little changed for the coefficients of transplantation factors and recipient characters. Across all time points (17 years times 12 months), the AUC value differed negligibly between the two models, with min, max and mean difference of 0.001, 0.006 and 0.002, respectively. The AUC value indicated both models had a moderate level of predictive power (Figure 1).

**Table 1.** Baseline characteristics of Black vs. Other donors.

| **Donor characteristics** | **Black**<br>(n=16,417) | **Other**<br>(n=103,024) | **P-value** |
| --- | --- | --- | --- |
| **Variable** | **Counts (%)** | **Counts (%)** |  |
| Gender (Male) | 9,834 (59.90) | 61,540 (59.73) | 0.68 |
| Cause of death: CVA | 7,003 (42.66) | 37,804 (36.69) | <0.001 |
| Diabetic | 1,440 (8.77) | 7,163 (6.95) | <0.001 |
| Hypertensive | 6,176 (37.62) | 28,284 (27.45) | <0.001 |
| Donation after cardiac death | 1,249 (7.61) | 16,330 (15.85) | <0.001 |
| Positive for Hepatitis C virus | 377 (2.30) | 3,021 (2.93) | <0.001 |
|  | **Mean (SD)** | **Mean (SD)** |  |
| Age | 36.17 (17.02) | 39.32 (16.45) | <0.001 |
| Height | 167.21 (22.14) | 168.70 (18.15) | 0.16 |
| Weight | 79.27 (27.84) | 79.33 (24.25) | 0.09 |
| Serum creatinine | 1.32 (1.05) | 1.14 (0.96) | <0.001 |

CVA = cerebrovascular accident; SD = standard deviation

**Figure 1.**
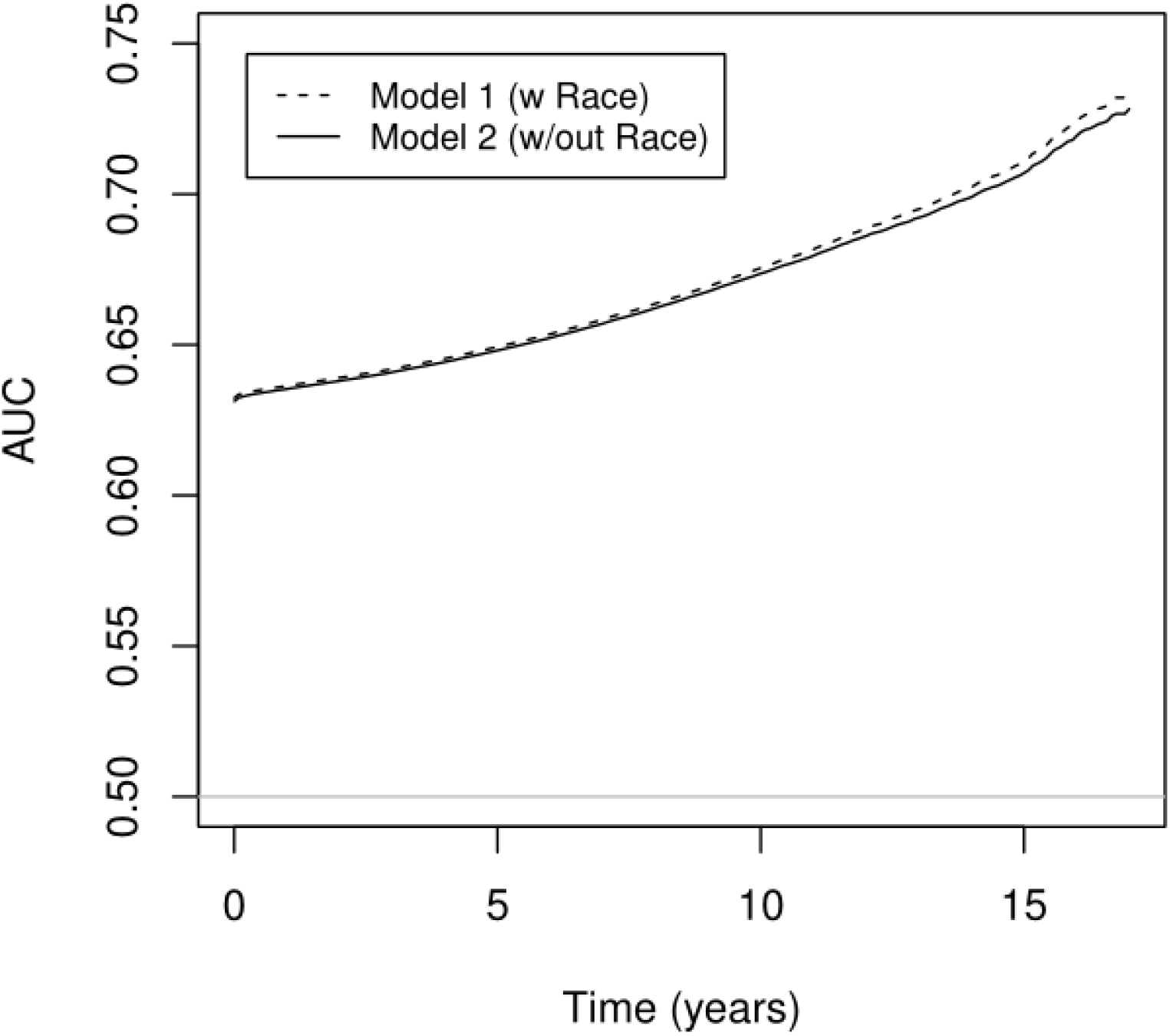
Time dependent AUC of models with or without donor race.

Our modeling and analysis showed that removing donor race from the original KDRI did not alter the overall model predictability of allograft failure or patient mortality. Clinical risk factors included in the KDRI have largely accounted for differential risk between black and other donors. Adding donor race into the KDRI only shifts how risk is attributed to these clinical risk factors, without yielding better prediction of outcomes than the model without race. Therefore, the effect of black donor race on the survival outcome beyond these risk factors is negligible and its inclusion in the KDRI is unwarranted. Our analysis suggests if race is used as a proxy of biological risk factors, its rationale should be supported by sufficient evidence that is appropriate and relevant. Precision medicine research offers the future promise to identify genetic, environmental, and lifestyle factors that will guide decision-making in areas such as kidney transplantation. Even if donor race were a proxy for any heretofore unknown but relevant biological factors, such as the APOL1 gene variants, it is statistically more valid and scientifically more rigorous to incorporate the APOL1 genotype, rather than race, in the calculation of risk for allograft failure.^10^ Finally, social determinants of health gleaned from precision medicine could, and should, replace race when accounting for the effects of disparities on transplant outcomes.

## Data Availability

Data are available through the United States Organ Sharing System

https://unos.org/

## Author’s Contribution

Research idea and study design: YZ, CA and YN; statistical analysis: IL and YZ; data analysis/interpretation/drafting and revision of manuscript: KC, IL, SC, YZ, CA and YN; Each author contributed important intellectual content during manuscript drafting or revision and agrees to be personally accountable for the individual’s own contributions and to ensure that questions pertaining to the accuracy or integrity of any portion of the work, even one in which the author was not directly involved, are appropriately investigated and resolved, including with documentation in the literature if appropriate.

## Supplement material

**Model Comparison: Survival model with 119,441 recipients and 38,804 events**

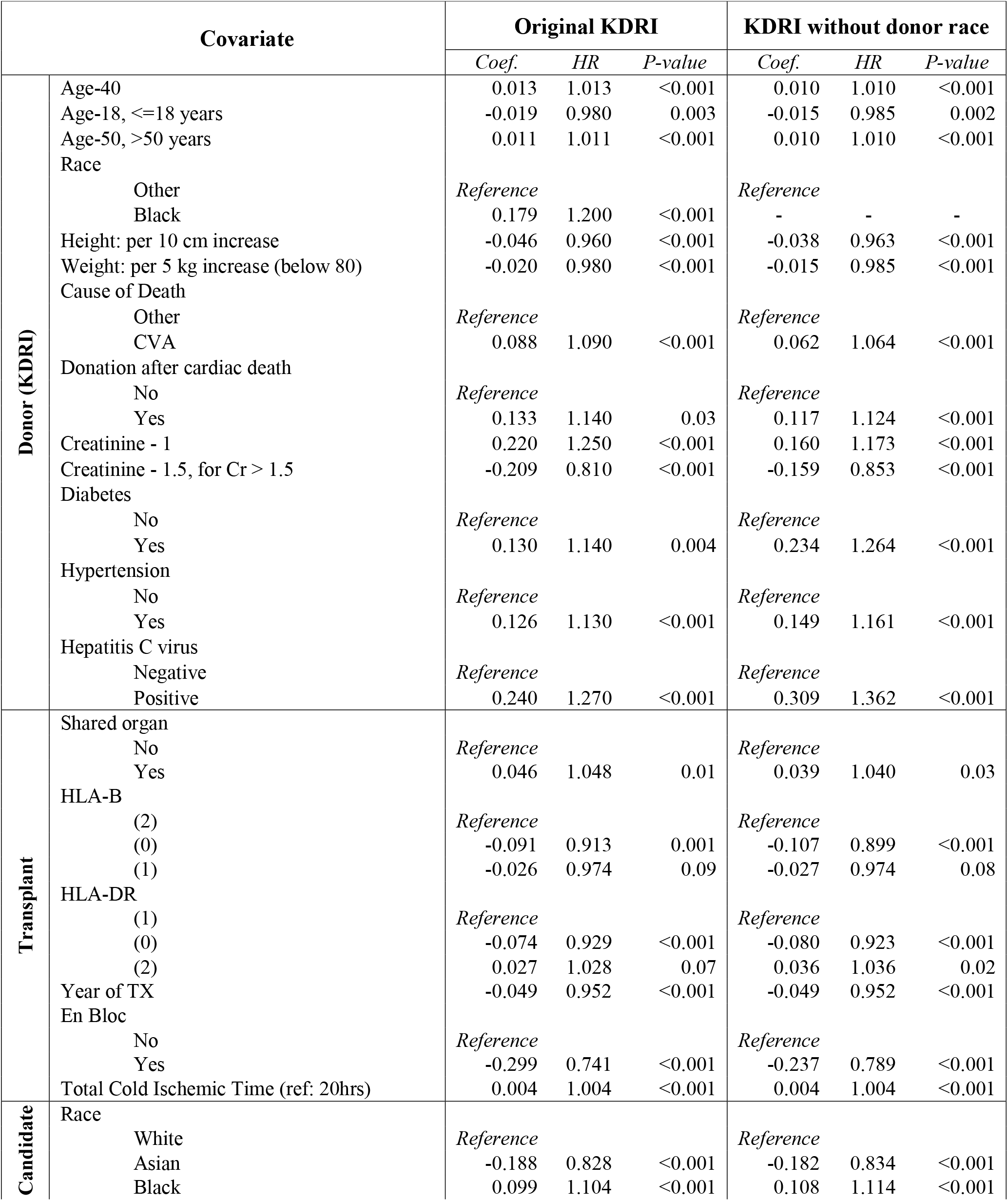
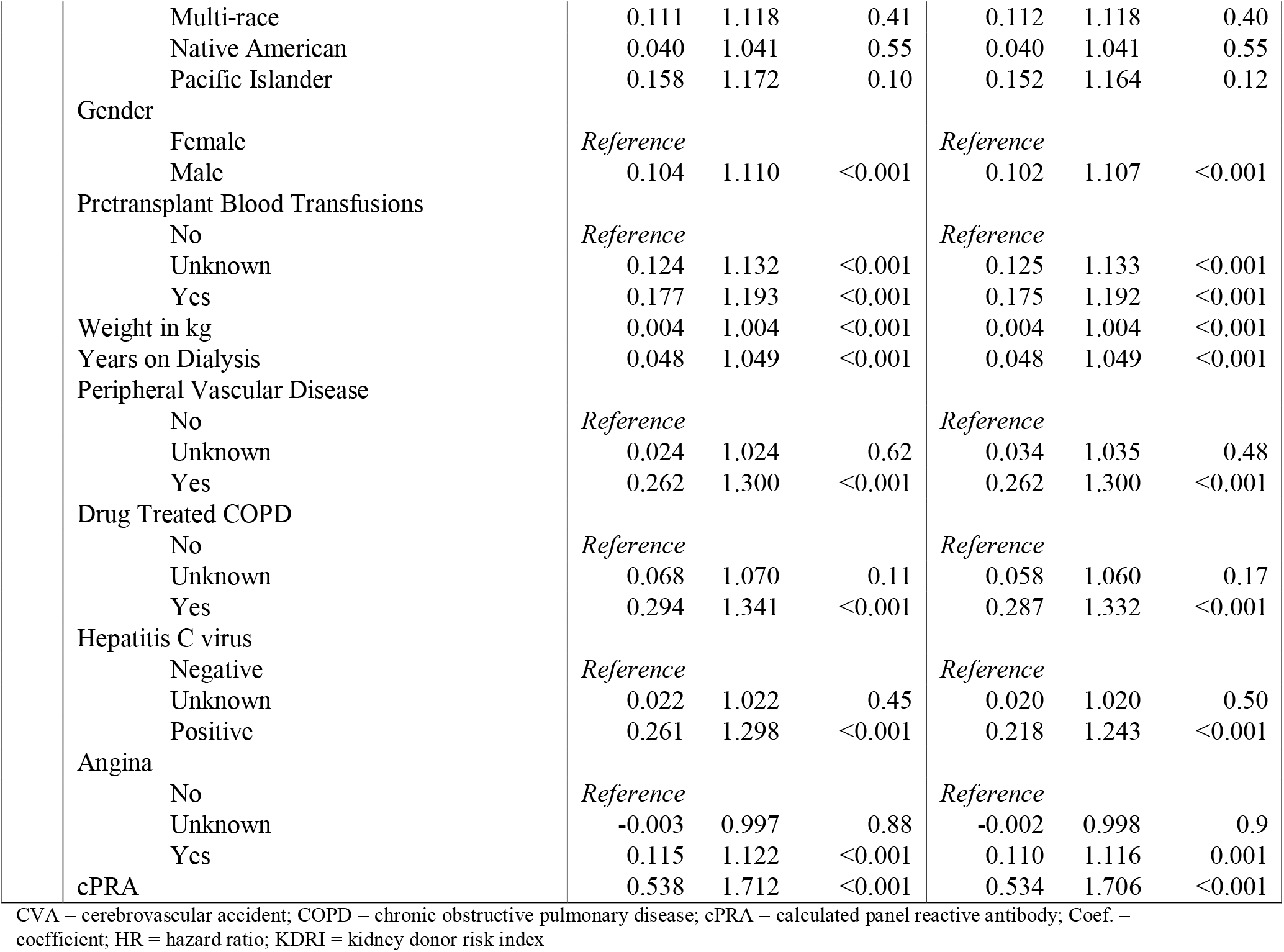

## References

1. Fontanarosa PB, Bauchner H. Race, Ancestry, and Medical Research. Jama. 2018;320(15):1539–1540.

2. Eneanya ND, Yang W, Reese PP. Reconsidering the Consequences of Using Race to Estimate Kidney Function. Jama. 2019;322(2):113–114.

3. Grubbs V. Precision in GFR Reporting: Let’s Stop Playing the Race Card. Clin J Am Soc Nephrol. 2020;15(8):1201–1202.

4. Hsu J, Johansen KL, Hsu CY, Kaysen GA, Chertow GM. Higher serum creatinine concentrations in black patients with chronic kidney disease: beyond nutritional status and body composition. Clin J Am Soc Nephrol. 2008;3(4):992–997.

5. Swanson SJ, Hypolite IO, Agodoa LY, et al. Effect of donor factors on early graft survival in adult cadaveric renal transplantation. American journal of transplantation: official journal of the American Society of Transplantation and the American Society of Transplant Surgeons. 2002;2(1):68–75.

6. Freedman BI, Julian BA, Pastan SO, et al. Apolipoprotein L1 gene variants in deceased organ donors are associated with renal allograft failure. American journal of transplantation: official journal of the American Society of Transplantation and the American Society of Transplant Surgeons. 2015;15(6):1615–1622.

7. Rao PS, Schaubel DE, Guidinger MK, et al. A comprehensive risk quantification score for deceased donor kidneys: the kidney donor risk index. Transplantation. 2009;88(2):231–236.

8. Package ‘survAUC’. https://cran.r-project.org/web/packages/survAUC/survAUC.pdf.

9. Chambless LE, Diao G. Estimation of time-dependent area under the ROC curve for long-term risk prediction. Statistics in medicine. 2006;25(20):3474–3486.

10. Julian BA, Gaston RS, Brown WM, et al. Effect of Replacing Race With Apolipoprotein L1 Genotype in Calculation of Kidney Donor Risk Index. American journal of transplantation: official journal of the American Society of Transplantation and the American Society of Transplant Surgeons. 2017;17(6):1540–1548.

